# Global, Regional, and National Burdens of Ischemic Stroke Attributed to High Body-mass Index from 1990 to 2021

**DOI:** 10.1101/2024.10.20.24315842

**Authors:** Yuan Zhang, Xuesong Ren, Xiaofeng Zhao, Lina Meng, Hai Lu, Chunhong Zhang

**Affiliations:** Department of Acupuncture and Moxibustion, First Teaching Hospital of Tianjin University of Traditional Chinese Medicine, Tianjin 300380, China; Graduate College, Tianjin University of Traditional Chinese Medicine, Tianjin 301600, China; National Clinical Research Center for Chinese Medicine Acupuncture and Moxibustion, Tianjin 300380, China; Department of Traditional Chinese Medicine, Nanjing Drum Tower Hospital, Affiliated Hospital of Medical School, Nanjing University, Nanjing 210003, China

**Keywords:** high body-mass index, ischemic stroke, global burden of disease, obesity, epidemiology

## Abstract

**Background:** Ischemic stroke (IS) is a significant contributor to the global disease burden, with overweight/obesity recognized as one of the primary risk factors. This study aims to evaluate the distribution and changes in the global burden of IS attributable to high body-mass index (BMI) from 1990 to 2021, providing a foundation for the development of targeted prevention and treatment strategies.

**Methods:** Based on the modeling strategies of the 2021 global burden of disease (GBD) dataset, the burden of IS attributable to high BMI was estimated. The assessment covered the global disease burden and its distribution by region, country, development level, age group, and gender, as well as temporal trends. Age-standardized rates, along with the estimated annual percentage change (EAPC) in these rates, were utilized to describe this burden and its evolving trends. Locally estimated scatterplot smoothing (LOESS) regression was employed to analyze the trends of disease burden in relation to changes in socio-demographic index (SDI).

**Results:** From 1990 to 2021, although the age-standardized death rate and disability-adjusted life years (DALY) rate for IS attributable to high BMI have declined globally, the absolute number of deaths and DALYs nearly doubled, reaching 172,390.65 (95% UI, 347,912.75 to 25,064.57) and 4,439,186.31 (95% UI, 8,647,485.11 to 649,030.21) respectively. Meanwhile, significant disparities in disease burden existed across different regions, genders, and age groups. In high-middle SDI regions, the burden of IS caused by high BMI significantly exceeded that in other regions, while in low-income areas, this burden was rapidly increasing. Additionally, the increase in burden among men was significantly greater than that among women, and the previously concentrated burden in the elderly is now rapidly shifting toward younger groups.

**Conclusions:** The overweight and obese populations continue to present significant challenges in managing the burden of IS. In light of future population growth and aging trends, more effective stroke prevention strategies and practical solutions are urgently needed, particularly in low- and middle-income countries. Additionally, given the increasing disease burden among men and younger populations, it is essential to establish targeted preventive measures to address the specific needs of different groups and improve overall health outcomes.

## 1 Introduction

According to the 2021 global burden of disease (GBD) study, stroke remained one of the leading causes of global disease burden, ranking as the third most common cause of death worldwide and the third leading non-communicable disease contributing to disability-adjusted life years (DALYs).^1,2^ Ischemic stroke (IS) accounted for approximately 87% of all strokes.^3^ Although the age-standardized incidence, death, and DALY rates for IS have decreased over the past 30 years, the absolute numbers of new cases, deaths, and DALYs have significantly increased due to global population growth and aging, with this trend expected to continue.^4–6^ Furthermore, IS imposed a substantial economic burden on society. In 2019, the value of lost welfare due to IS reached $964.51 billion, constituting 0.78% of global gross domestic product, with significant disparities across different regions and socioeconomic groups.^7,8^ Therefore, effectively preventing and controlling IS has become a major challenge in global public health.

As society develops and lifestyles change, metabolic risk has become the leading level 1 GBD risk factor for stroke.^2^ Among these risks, high body-mass index (BMI) is one of the most common, associated with various non-communicable diseases.^2,9^ BMI, calculated as weight (kg) divided by height (m) squared, is the most commonly used measure for assessing overweight and obesity.^10,11^ As the second largest preventable causes of death (after tobacco) globally, overweight and obesity have become increasingly severe and represent a significant public health concern.^12^ Research indicated that an excessively high BMI can trigger metabolic disorders, such as elevated blood pressure, dyslipidemia, and insulin resistance, while also exacerbating chronic systemic inflammation, which contributes to atherosclerosis and ultimately increases the risk of IS.^12–14^ For individuals with a BMI over 20, the risk of IS increases by 5% for each additional BMI unit.^15,16^ Similarly, regarding stroke mortality, a gradual and direct dose-response relationship was observed when BMI exceeded 25, whereas no significant association was seen for BMI below 25.^17^ In recent years, high BMI has become the fastest-growing risk factor for age-standardized DALYs related to IS worldwide, showing a concerning upward trend.^2^ Therefore, assessing the global burden of IS attributable to high BMI has become critically important.

In this study, we estimated the global burden trends of high BMI-related IS from 1990 to 2021, analyzed variations across different countries and regions, and systematically summarized the high-risk populations based on gender, age, and socio-demographic index (SDI), aiming to provide a foundation for targeted prevention and intervention measures.

## 2 Methods

### 2.1 Data sources

The data used in this study to estimate the burden of IS attributable to high BMI were derived from the GBD 2021 dataset, accessible via the GBD Results Tool on the Global Health Data Exchange platform (https://vizhub.healthdata.org/gbd-results/). The GBD 2021 database provides information on mortality and health loss caused by 288 causes of death, 371 diseases and injuries, and 88 risk factors across 204 countries and territories from 1990 to 2021. This study followed the Guidelines for Accurate and Transparent Health Estimates Reporting (GATHER).^18^ Based on data availability, we extracted the number and rate of deaths and DALYs due to IS attributable to high BMI, stratified by year, age, sex, and region.

### 2.2 Definitions

In GBD 2021 study, the average BMI was calculated using weight and height information from available individual-level survey data.^19^ High BMI for adults is defined as a BMI greater than 20 to 23 kg/m². This theoretical minimum risk exposure level (TMREL) was determined based on the BMI level associated with the lowest risk of all-cause mortality.^19,20^ IS is characterized by the occlusion of blood flow to parts of the brain due to thrombosis or embolism, leading to ischemia and hypoxia in the affected brain tissue, ultimately resulting in neurological dysfunction. According to the world health organization (WHO) criteria, IS is defined as rapidly developing clinical signs of (usually focal) disturbance of cerebral function lasting more than 24 hours or leading to death.^21,22^ In GBD 2021, IS encompassed several disease categories identified by ICD-10 codes, including G45-G46.8, I63-I63.9, I65-I66.9, I67.2-I67.3, I67.5-I67.6, and I69.3.^23^

The geospatial scope used in this study was divided into four levels: global, 5 SDI regions, 21 super-regions with epidemiological similarities and geographic proximity, and 204 individual countries or territories. The SDI, developed by GBD researchers, is a composite indicator reflecting the development levels closely associated with health outcomes across countries and regions. It is the geometric mean of 0 to 1 indices of total fertility rate under the age of 25, mean education for those ages 15 and older, and lag distributed income per capita.^24^ In 2021, the SDI classification was as follows: low (<0.47), low-middle (0.47–0.62), middle (0.62–0.71), high-middle (0.71–0.81), and high (>0.81).^24^

### 2.3 Burden Estimation

Based on the comparative risk assessment framework, the GBD 2021 risk factor analysis estimated the burden of deaths and DALYs associated with IS attributable to high BMI at global, regional, and national levels.^25^ Specifically, the GBD primarily used a meta-regression in the burden of proof approach to identify and extract data from published systematic reviews, enabling the estimation of relative risks for risk-outcome pairs.^26^ On this basis, bayesian statistical model (spatio-temporal Gaussian process regression) was used to estimate the prevalence of overweight and obesity. The non-linear dose-response relationship between high BMI and disease endpoints was then modeled using meta-regression—Bayesian, regularized, trimmed. After estimating the all-age, cause-specific dose-response risk curves, the TMREL was determined. Population-attributable fractions were then calculated to measure the attributable burden. Detailed methods, and definitions of technical terms have been described elsewhere.^1,19^

### 2.4 Statistical Analysis

Between 1990 and 2021, the global burden estimates of IS attributable to high BMI were given as death and DALY rates per 100,000 population. To ensure comparability, the data were standardized using the GBD standard population structure. All estimates were computed based on the mean of 500 draws, with the 95% uncertainty intervals (UI) determined by the 2.5th and 97.5th percentiles of the distribution.

The estimated annual percentage change (EAPC) was used to evaluate trends in age-standardized rates (ASR) from 1990 to 2021. The EAPC and 95% confidence intervals (CI) were calculated using log-linear regression. Specifically, a linear regression model was fitted between time (year) and the log-transformed ASR: ln(ASR)=α+βx+ε, where x represents the year, and β is the regression coefficient. The EAPC was then calculated as 100×(exp[β]−1), with the 95% CI determined based on the standard errors from the log-linear regression.^27^ If the lower limit of the EAPC’s 95% CI was >0, the ASRs were considered to be increasing over the observation period. Conversely, the ASRs were considered to show a decreasing trend if the upper limit of the EAPC’s 95% CI was <0. When the 95% CI included both positive and negative values, the ASRs were considered relatively stable. Locally estimated scatterplot smoothing (LOESS) regression was used to analyze the trend of disease burden changes relative to the SDI. All tests were two-sided, and a p-value of <0.05 was considered statistically significant. Statistical analyses were performed using R software (v.4.3.3, http://www.r-project.org/).

## 3 Results

### 3.1 Global Burden of IS Attributable to High BMI

In 2021, high BMI-related IS resulted in 172,390.65 (95% UI, 347,912.75 to 25,064.57) deaths and 4,439,186.31 (95% UI, 8,647,485.11 to 649,030.21) DALYs globally. Compared to 1990, these numbers have approximately doubled. However, after further adjusting for time and age structure, the burden showed a relative decrease. The age-standardized death rate decreased from 2.57 (95% UI, 5.15 to 0.37) per 100,000 population in 1990 to 2.06 (95% UI, 4.17 to 0.30) in 2021, with an EAPC of -1.10 (95% CI, -0.96 to -1.25). Similarly, the age-standardized DALY rate decreased from 55.30 (95% UI, 109.16 to 7.90) per 100,000 population in 1990 to 51.52 (95% UI, 100.28 to 7.52) in 2021, with an EAPC of -0.58 (95% CI, -0.46 to -0.71). Detailed information is presented in Table 1.

**Table 1.**
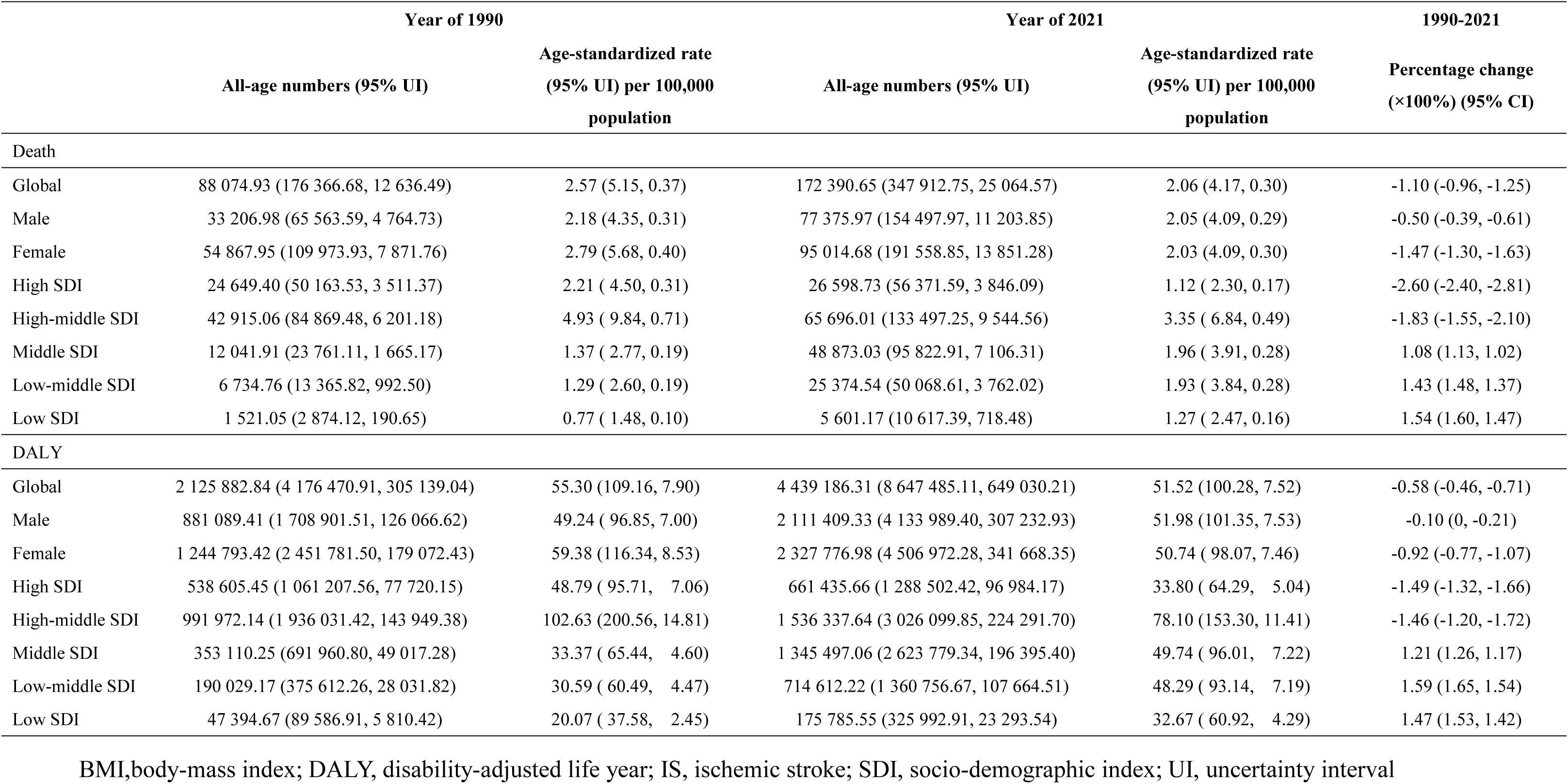
Burden of IS Attributable to High BMI by Sex and SDI Regions in 1990 and 2021.

|  | Year of 1990 |  | Year of 2021 |  | 1990-2021 |
| --- | --- | --- | --- | --- | --- |
|  | All-age numbers (95% UI) | Age-standardized rate<br>(95% UI) per 100,000<br>population | All-age numbers (95% UI) | Age-standardized rate<br>(95% UI) per 100,000<br>population | Percentage change<br>(×100%) (95% CI) |
| Death |  |  |  |  |  |
| Global | 88 074.93 (176 366.68, 12 636.49) | 2.57 (5.15, 0.37) | 172 390.65 (347 912.75, 25 064.57) | 2.06 (4.17, 0.30) | -1.10 (-0.96, -1.25) |
| Male | 33 206.98 (65 563.59, 4 764.73) | 2.18 (4.35, 0.31) | 77 375.97 (154 497.97, 11 203.85) | 2.05 (4.09, 0.29) | -0.50 (-0.39, -0.61) |
| Female | 54 867.95 (109 973.93, 7 871.76) | 2.79 (5.68, 0.40) | 95 014.68 (191 558.85, 13 851.28) | 2.03 (4.09, 0.30) | -1.47 (-1.30, -1.63) |
| High SDI | 24 649.40 (50 163.53, 3 511.37) | 2.21 ( 4.50, 0.31) | 26 598.73 (56 371.59, 3 846.09) | 1.12 ( 2.30, 0.17) | -2.60 (-2.40, -2.81) |
| High-middle SDI | 42 915.06 (84 869.48, 6 201.18) | 4.93 ( 9.84, 0.71) | 65 696.01 (133 497.25, 9 544.56) | 3.35 ( 6.84, 0.49) | -1.83 (-1.55, -2.10) |
| Middle SDI | 12 041.91 (23 761.11, 1 665.17) | 1.37 ( 2.77, 0.19) | 48 873.03 (95 822.91, 7 106.31) | 1.96 ( 3.91, 0.28) | 1.08 (1.13, 1.02) |
| Low-middle SDI | 6 734.76 (13 365.82, 992.50) | 1.29 ( 2.60, 0.19) | 25 374.54 (50 068.61, 3 762.02) | 1.93 ( 3.84, 0.28) | 1.43 (1.48, 1.37) |
| Low SDI | 1 521.05 (2 874.12, 190.65) | 0.77 ( 1.48, 0.10) | 5 601.17 (10 617.39, 718.48) | 1.27 ( 2.47, 0.16) | 1.54 (1.60, 1.47) |
| DALY |  |  |  |  |  |
| Global | 2 125 882.84 (4 176 470.91, 305 139.04) | 55.30 (109.16, 7.90) | 4 439 186.31 (8 647 485.11, 649 030.21) | 51.52 (100.28, 7.52) | -0.58 (-0.46, -0.71) |
| Male | 881 089.41 (1 708 901.51, 126 066.62) | 49.24 ( 96.85, 7.00) | 2 111 409.33 (4 133 989.40, 307 232.93) | 51.98 (101.35, 7.53) | -0.10 (0, -0.21) |
| Female | 1 244 793.42 (2 451 781.50, 179 072.43) | 59.38 (116.34, 8.53) | 2 327 776.98 (4 506 972.28, 341 668.35) | 50.74 ( 98.07, 7.46) | -0.92 (-0.77, -1.07) |
| High SDI | 538 605.45 (1 061 207.56, 77 720.15) | 48.79 ( 95.71, 7.06) | 661 435.66 (1 288 502.42, 96 984.17) | 33.80 ( 64.29, 5.04) | -1.49 (-1.32, -1.66) |
| High-middle SDI | 991 972.14 (1 936 031.42, 143 949.38) | 102.63 (200.56, 14.81) | 1 536 337.64 (3 026 099.85, 224 291.70) | 78.10 (153.30, 11.41) | -1.46 (-1.20, -1.72) |
| Middle SDI | 353 110.25 (691 960.80, 49 017.28) | 33.37 ( 65.44, 4.60) | 1 345 497.06 (2 623 779.34, 196 395.40) | 49.74 ( 96.01, 7.22) | 1.21 (1.26, 1.17) |
| Low-middle SDI | 190 029.17 (375 612.26, 28 031.82) | 30.59 ( 60.49, 4.47) | 714 612.22 (1 360 756.67, 107 664.51) | 48.29 ( 93.14, 7.19) | 1.59 (1.65, 1.54) |
| Low SDI | 47 394.67 (89 586.91, 5 810.42) | 20.07 ( 37.58, 2.45) | 175 785.55 (325 992.91, 23 293.54) | 32.67 ( 60.92, 4.29) | 1.47 (1.53, 1.42) |

BMI,body-mass index; DALY, disability-adjusted life year; IS, ischemic stroke; SDI, socio-demographic index; UI, uncertainty interval

### 3.2 Burden of IS Attributable to High BMI by Region and Country

In 2021, the burden of IS attributable to high BMI varied across the 21 super regions (Supplemental Table 1). The regions with the highest burden included Eastern Europe, North Africa and the Middle East, and Central Asia, where the age-standardized death rates > 5 per 100,000 population, and the age-standardized DALY rates > 125 per 100,000 population. In contrast, the region with the lowest age-standardized death and DALY rates was High-income Asia Pacific, with rates of 0.34 (95% UI, 0.68 to 0.05) and 11.47 (95% UI, 22.05 to 1.55), respectively. From 1990 to 2021, 10 out of the 21 super regions experienced an upward trend in age-standardized death and DALY rates due to high BMI-related IS. The most significant increases were observed in Asian regions, including East Asia, South Asia, and Southeast Asia, where the EAPCs for both death and DALY rates > 2.5. The most notable decline occurred in Western Europe, with EAPCs of -3.68 (95% CI, -3.52 to -3.84) for death rates and -2.91 (95% CI, -2.75 to -3.08) for DALY rates.

At the national level, the disparities in disease burden were even more significant. (Supplemental Table 2 and Figure 1). In 2021, Egypt had the highest age-standardized death rate at 15.81 (95% UI, 31.83 to 2.58) per 100,000 population, while Singapore had the lowest at 0.21 (95% UI, 0.44 to 0.03), a difference of up to 75 times. From 1990 to 2021, 107 countries or regions experienced an upward trend in the burden of IS due to high BMI. Notably, Zimbabwe, Lesotho, Indonesia, and Vietnam had EAPCs for age-standardized death rates all > 4. Conversely, 86 countries or regions declined in disease burden, with Estonia, the Czech Republic, Austria, and Portugal showing the most significant decreases, with EAPCs for age-standardized death rates all ≤ -5. The changes in age-standardized DALY rates followed a similar pattern to death rates.

**Figure 1.**
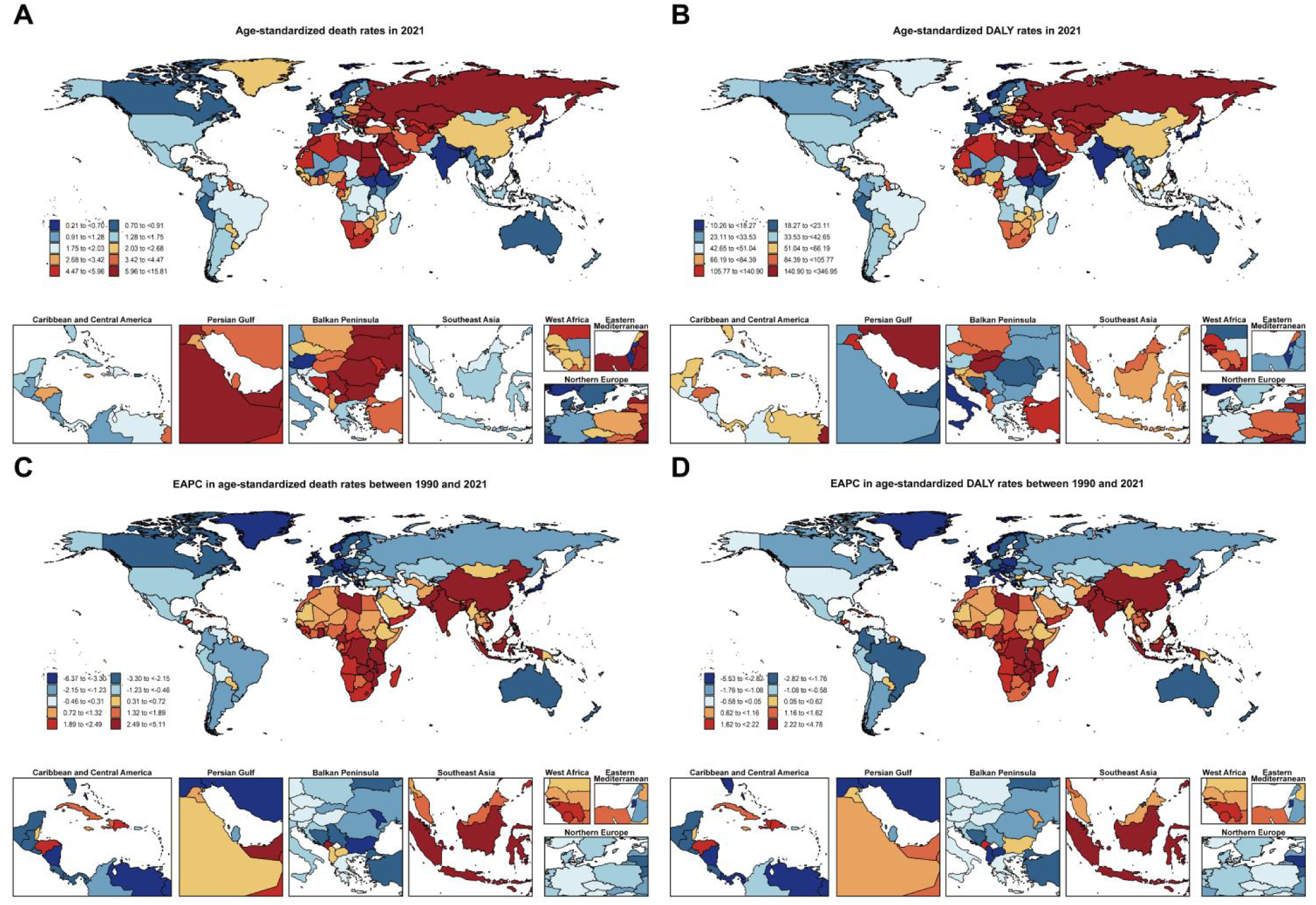
Global Maps of the Burden of IS Attributable to High BMI Abbreviations: BMI, body-mass index; DALY, disability-adjusted life year; EAPC, estimated annual percentage change; IS, ischemic stroke.

### 3.3 Burden of IS Attributable to High BMI by SDI Region

Globally, the disease burden across different SDI regions has shown uneven development trends (Table 1). In the past 31 years, high SDI regions have experienced the most significant decline in the burden of IS attributable to high BMI, with the age-standardized death rates in 2021 being the lowest among all SDI regions at 1.12 (95% UI, 2.30 to 0.17). Meanwhile, although the disease burden in high-middle SDI regions has also decreased, their age-standardized death rates in 2021 remained significantly higher than other SDI regions, reaching 3.35 (95% UI, 6.84 to 0.49). From 1990 to 2021, the burden of IS due to high BMI increased in middle, low-middle, and low SDI regions, with the fastest growth in age-standardized death rates observed in low SDI regions at 1.54 (95% CI, 1.60 to 1.47), and the highest increase in age-standardized DALY rates occurred in low-middle SDI regions at 1.59 (95% CI, 1.65 to 1.54).

Figure 2 illustrates the relationship between SDI levels across various super regions and the age-standardized death and DALY rates of IS attributable to high BMI. To capture potential nonlinear relationships, LOESS regression was employed. The LOESS curve reveals a turning point at an SDI value of 0.7: when SDI < 0.7, the age-standardized death and DALY rates gradually increase with rising SDI; however, once SDI > 0.7, both rates decrease rapidly as SDI continues to grow.

**Figure 2.**
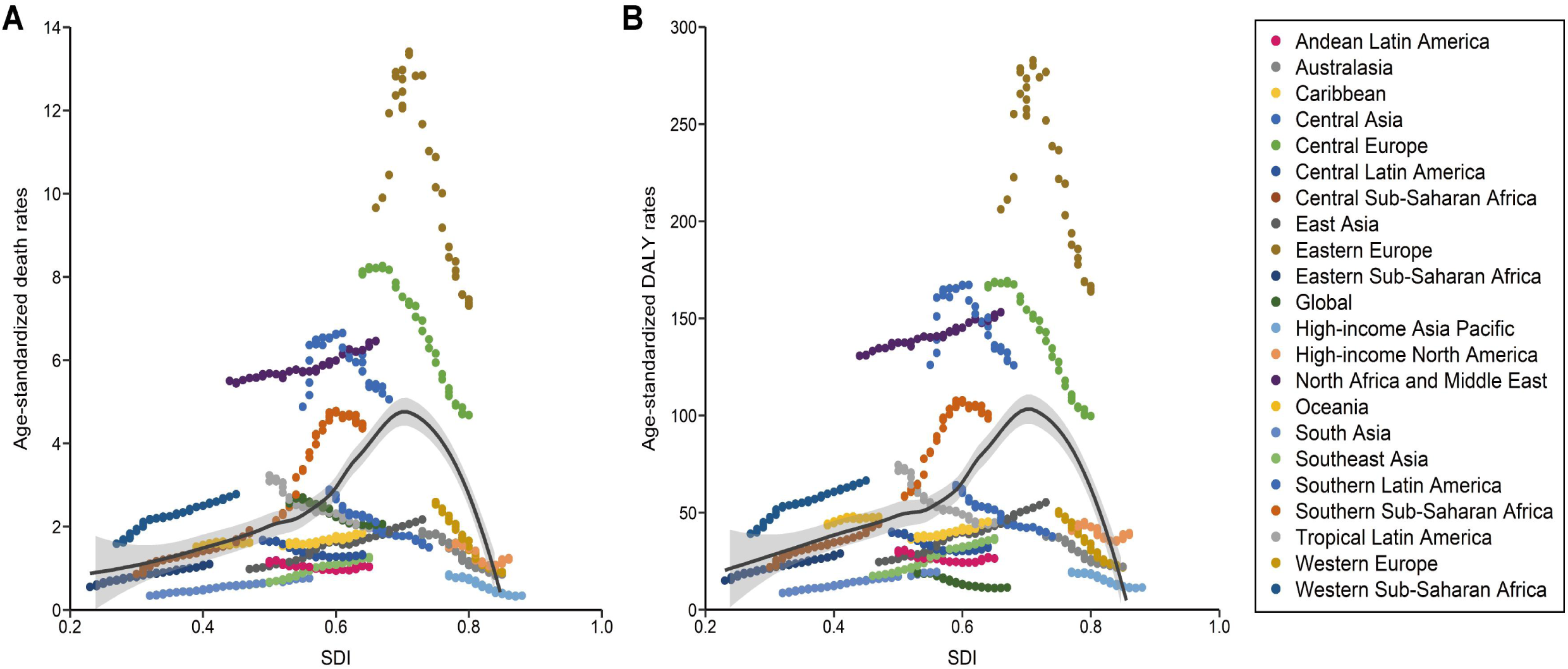
Global Burden of IS Attributable to High BMI with SDI, by Region from 1990 to 2021 Abbreviations: BMI, body-mass index; DALY, disability-adjusted life year; IS, ischemic stroke; SDI, socio-demographic index.

### 3.4 Burden of IS Attributable to High BMI by Sex and Age

There was a sex-related disparity in the burden of IS attributable to high BMI (Table 1). From 1990 to 2021, both men and women experienced a decline in IS burden due to high BMI, with the rate of decline being markedly greater for women. Specifically, the EAPC in age-standardized DALY rates were -0.1 (95% CI, 0 to -0.21) for men and -0.92 (95% CI, -0.77 to -1.07) for women, showing a ninefold difference. This led to a shift from a higher disease burden in women compared to men in 1990, to a higher burden in men by 2021. Further analysis across different SDI regions revealed additional gender differences in burden (Figure 3). From 1990 to 2021, both men and women in high and high-middle SDI regions saw a decline in IS burden due to high BMI, with women experiencing a greater reduction. However, in middle, low-middle, and low SDI regions, the burden increased, with men showing a higher rate of increase.

**Figure 3.**
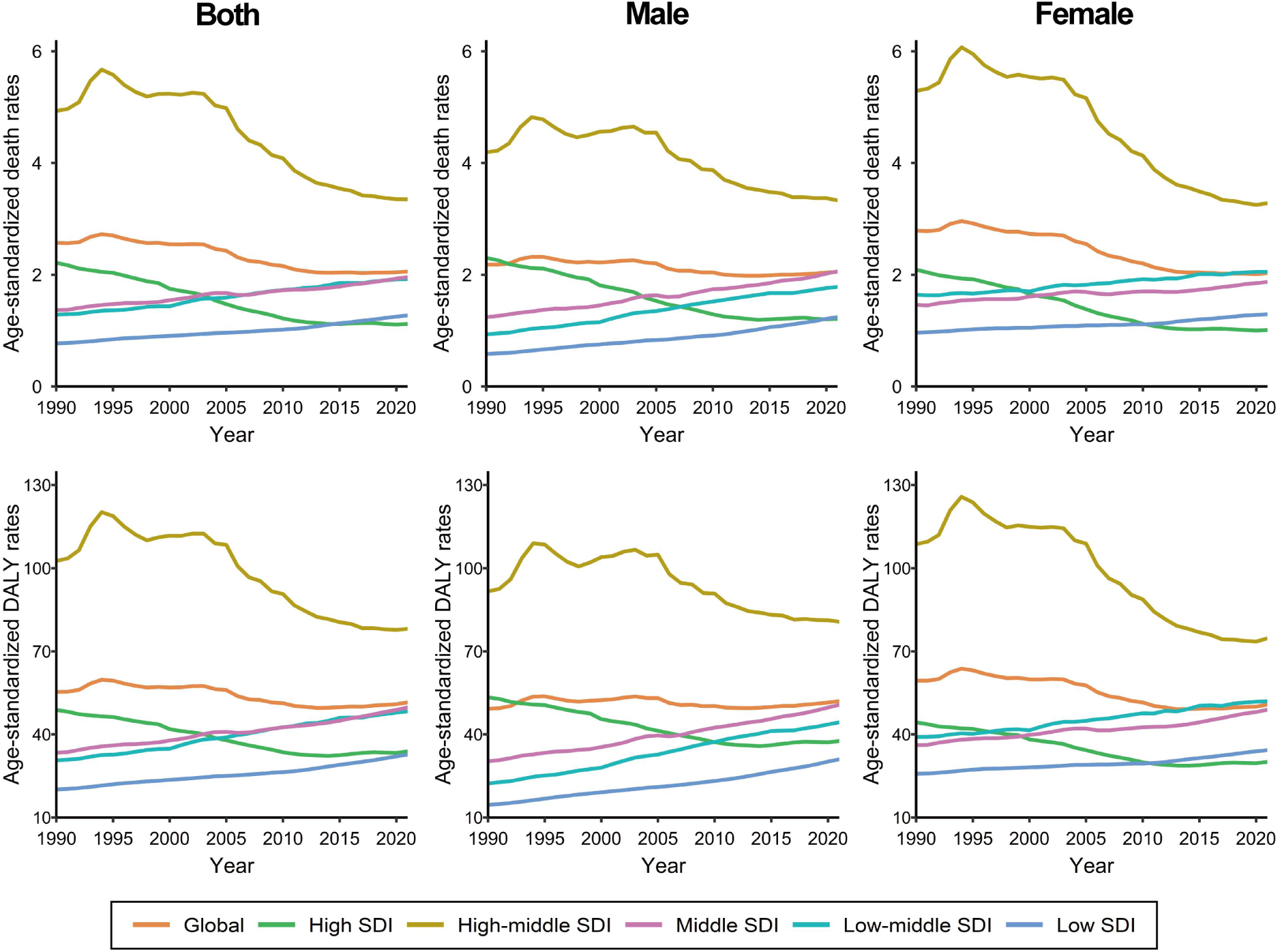
Trend of Global Burden of IS Attributable to High BMI by Sex and SDI Regions from 1990 to 2021 Abbreviations: BMI, body-mass index; DALY, disability-adjusted life year; IS, ischemic stroke; SDI, socio-demographic index.

There are significant differences in the burden of IS attributable to high BMI across different age groups (Supplemental Table 3). In 2021, the burden of IS due to high BMI increased markedly with age, peaking in individuals aged 80 and older, with a death rate of 35.73 (95% UI, 79.91 to 4.76). Conversely, the burden was lowest in the 20-24 age group, where the death rate was only 0.02 (95% UI, 0.05 to 0.00). Further analysis by gender within each age group revealed the following trends: among individuals aged 20-24 and those aged 70 and above, women exhibited higher death and DALY rates compared to men. In age 25-44, although women had lower death rates than men, their DALY rates were higher. For individuals aged 45-69, both death and DALY rates were greater for men (Supplemental Figure 2). From 1990 to 2021, both men and women aged 20-44 showed an upward trend in death and DALY rates, with younger individuals experiencing a more pronounced increase. While the burden for individuals aged 60 and older exhibited a declining trend, with the decrease becoming more rapid as age increased. Across all age groups, the EAPCs for death and DALY rates were consistently higher in men (Figure 4).

**Figure 4.**
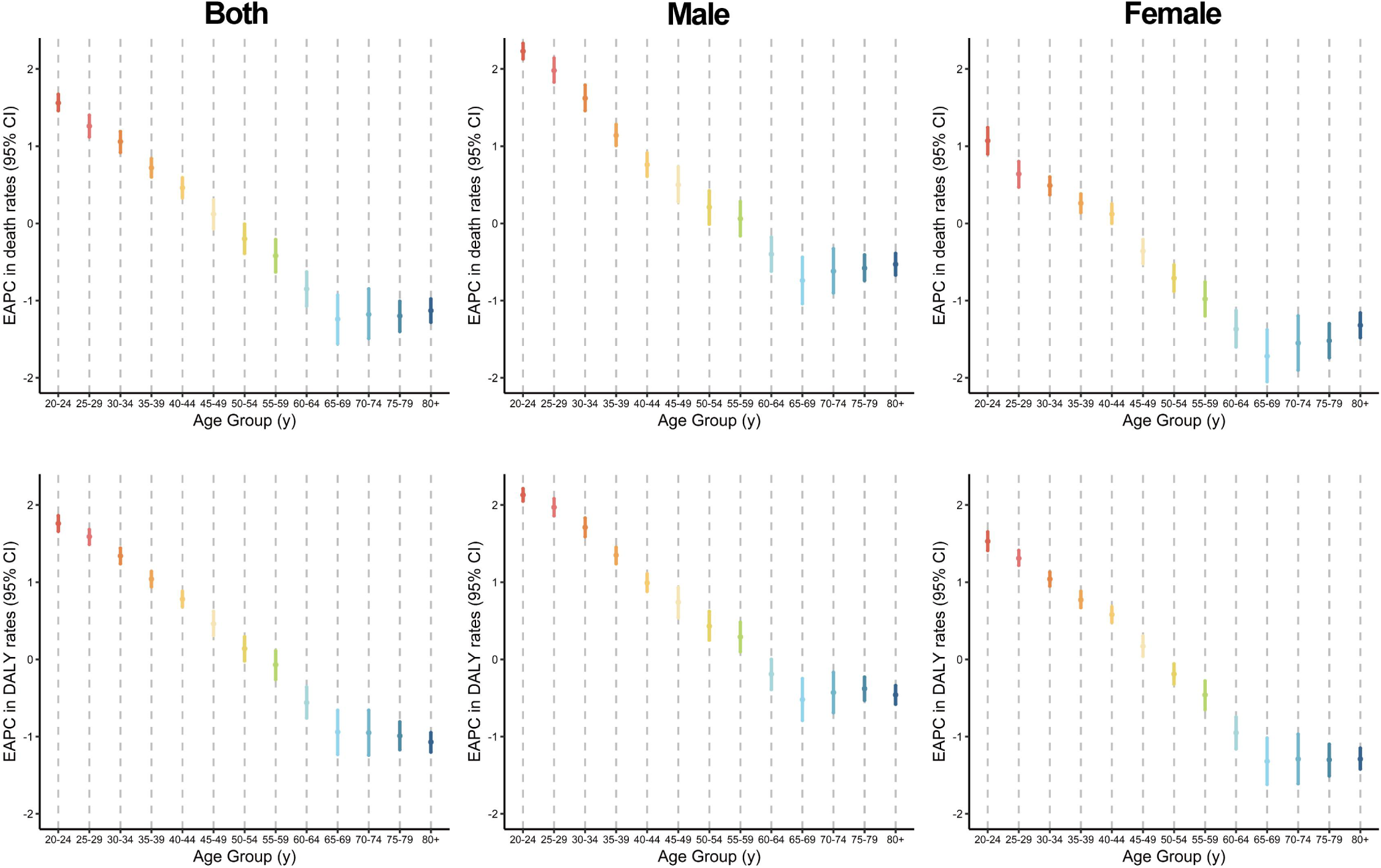
EAPCs in Age-Specific Death and DALY Rates of IS Attributable to High BMI by Age Group from 1990 to 2021 Abbreviations: BMI, body-mass index; DALY, disability-adjusted life year; EAPC, estimated annual percentage change; IS, ischemic stroke.

## 4 Discussion

This study is the first to assess the global burden of IS attributable to high BMI using the GBD 2021 dataset, revealing variations across different geographic regions, genders, and age groups. Overall, from 1990 to 2021, although the age-standardized death and DALY rates for IS due to high BMI have decreased globally, the absolute number of deaths and DALYs has doubled, suggesting that population growth and aging might be the main contributors to the increased burden. Additionally, significant differences in disease burden were observed across regions, genders, and age groups. In high-middle SDI regions, the burden of IS caused by high BMI was particularly severe, while in middle SDI, low-middle SDI, and low SDI regions, this burden was rapidly increasing. By 2021, the disease burden among men had surpassed that of women, however, among those under 45 and over 70, women still had higher DALY rates than men. Furthermore, although the elderly continued to bear a heavy disease burden, the IS burden caused by high BMI was gradually shifting toward younger populations.

High BMI is largely preventable, as evidenced by the global decline in IS burden attributed to high BMI, but the reduction has been significantly less than the overall decline in IS. Between 1990 and 2021, high BMI became the leading contributor to the increase in stroke-related DALYs among all level 4 risk factors (88.2% [53.4–117.7]).^2^ A noteworthy point is the significant socioeconomic disparity in the IS burden caused by high BMI: as the SDI increases, the initial disease burden rises but then declines markedly. This trend has been corroborated by other studies.^6^ Previous research indicated that BMI and the rates of overweight/obesity tended to increase steadily with economic development, for every 1% rise in income, the overweight/obesity rates increased by approximately 0.2-0.3%.^28–30^ However, the burden of IS attributable to high BMI was lowest in high-SDI regions but highest in high-middle SDI regions, which might be attributed to several factors. On one hand, high-SDI regions likely benefited from more advanced healthcare systems, effective public health interventions, and greater health awareness, these factors as well as other healthy behaviors might mitigate the negative impacts of obesity, while high-middle SDI regions faced lower healthcare quality, inadequate preventive measures, weaker health awareness, and a higher prevalence of comorbid chronic conditions, all of which intensified the relationship between high BMI and IS.^31–34^ On the other hand, there may be an "obesity paradox" in IS outcomes, where overweight patients (BMI 25–30) seem to have better post-stroke prognoses compared to those with normal weight (BMI 18.5–25), but this paradox may not apply to individuals with obesity (BMI > 30).^35,36^ Therefore, while high BMI was more prevalent in high-SDI regions, residents of these areas often maintained healthier diets and nutritional structures, which might result in overweight rather than obese status, thereby reducing some of the burden.^37,38^ Conversely, high-middle SDI regions often had diets high in salt, fat, and sugar, exacerbating obesity rates and increasing the related disease burden.^38–40^

As for middle SDI, low-middle SDI, and low SDI regions, although the prevalence of obesity and overweight is relatively low, the burden of IS caused by high BMI has been rising, particularly in parts of Africa and Asia. This trend is consistent with previous research, indicating that the impact of stroke has generally increased among lower-income populations, and this socioeconomic-driven negative effect may have a cumulative impact.^31,41,42^ Additionally, research has indicated that due to shifts in dietary patterns and rapid urbanization, these regions are increasingly facing a "double burden of malnutrition", where undernutrition and overnutrition coexist within the same population.^43,44^ Taking Southeast Asia as an example, over the past 30 years, overnutrition and undernutrition have become dual public health challenges.^45^ In this environment, local residents become more vulnerable to related diseases after gaining weight, particularly among children and adolescents, as increased BMI during this time may be directly linked to a higher risk of stroke in adulthood.^46^ Furthermore, due to inadequate medical resources and health management, people in these regions often cannot access timely interventions or preventive measures when cardiovascular problems arise from elevated BMI, making Southeast Asia one of the fastest-growing regions in terms of the burden of IS caused by high BMI between 1990 and 2021.^47^ Beyond these factors, the nutrition transition phenomenon, environment multiplier theory, and the thrifty gene hypothesis may also help explain the rapid rise in the high BMI-related IS burden in Southeast Asia.^48,49^

Regarding age, the burden of IS due to high BMI increased with age, peaking in death and DALY rates among individuals over 80. Age itself is an important and immutable prognostic risk factor for IS.^50^ The increased burden in older populations may be attributed to the cumulative effects of long-term unhealthy lifestyle factors such as sedentariness, heavy smoking, and excessive alcohol consumption, all of which contribute to metabolic syndrome and vascular diseases.^51,52^ Additionally, as people age, certain genetic factors may exert a greater impact on obesity and IS prognosis.^50^ Interestingly, while the burden intensified with age, this increase was more pronounced up to the age of 70 and then slowed thereafter. This finding aligns with a previous NHANES study, which showed that overweight/obese individuals under 70 had a higher post-stroke mortality rate than their normal-weight peers, while the trend reversed after 70.^53^

Our study also found that over the past 30 years, the burden of high BMI-related IS has been gradually shifting toward younger age groups, with the youngest showing the fastest increase in death and DALY rates. Compared to the elderly, the causes of stroke in young adults are more diverse, with about one-third of cases classified as cryptogenic strokes.^54^ These strokes often have a profound impact on patients’ social interactions, daily activities, and career development.^55,56^ Moreover, strokes in younger individuals impose a significant economic burden. It has been estimated that by 2050, nearly half of stroke-related costs in the U.S. will be attributed to strokes in younger populations.^57^ Therefore, targeted interventions are urgently needed to mitigate the impact of the rising IS burden among younger individuals on both personal and societal levels.

Additionally, this study revealed that the global burden of IS attributable to high BMI decreased faster in women compared to men. We hypothesized that this trend may be closely related to economic development and urbanization. Although men had a higher overall disease burden than women as of 2021, women in low-middle SDI and low SDI regions still exhibited higher mortality and DALY rates compared to men. This discrepancy may be partially explained by differences in exposure to IS risk factors between men and women in higher-income regions, where women often adopt healthier lifestyles, while risk factors such as smoking and drinking are more prevalent among men.^5,58^ What’s more, studies have shown that in countries with higher socioeconomic status, the burden of overweight and obesity among men is increasing at a faster rate than in other regions.^59^

Since women tend to experience IS later in life than men, the gender differences in high BMI-related IS burden may also be partly attributed to age-related differences in modifiable risk factors, which cause the burden to change dynamically over time.^60^ Our results showed that for individuals under 45 and over 70, women had a higher DALY rate compared to men. For younger women, potential causes may include unique female-specific risks (such as pregnancy and contraceptive use) and certain genetic disorders.^61,62^ Research also indicated that post-stroke lifestyle and behaviors are significant factors for young female stroke survivors,^63^ with obesity during this stage greatly increasing the risk of early death.^64^ As women age, hormonal changes, particularly after menopause, can affect fat distribution and metabolism. The decline in estrogen may lead to increased abdominal fat, inflammation, and insulin resistance, all of which negatively affect cardiovascular health.^65,66^ This, in turn, reduces pre-stroke functional capacity and exacerbates the burden of chronic diseases in older women,^67^ contributing to more severe strokes, higher mortality rates, and poorer functional outcomes.^68,69^ Furthermore, men with cardiovascular diseases are more likely to die early from heart-related conditions (i.e., “competing causes of death”), which may partially explain the relatively higher IS risk and burden among elderly women with obesity.^70^

This study has several limitations. First, in some underdeveloped regions, the low coverage of disease reporting agencies leads to a lack of reliable epidemiological data, which may result in an underestimation of the true disease burden. Second, using BMI as an indicator of overweight/obesity has its inherent limitations. BMI cannot distinguish between fat and muscle, nor does it account for fat distribution, with visceral fat having a greater impact on IS than subcutaneous fat. As a result, the burden of IS associated with high BMI may deviate from the actual situation. Third, variations in BMI responses across different ethnic groups and regions may obscure disparities in disease burden among populations. For example, Asian populations tend to exhibit higher health risks at lower BMI levels, which might affect the analysis to varying degrees. Finally, in the 2021 GBD database, high BMI was considered a risk factor, rendering prevalence data unavailable, which means the reported disease burden in this study might be a conservative estimate.

In summary, this study estimated the global, regional, and national burden of IS attributable to high BMI and further analyzed the burden differences across socioeconomic statuses, age groups, and genders, which may help to more effectively identify high-risk populations. The results underscore the need to develop more effective stroke prevention strategies and pragmatic solutions in the context of future population growth and aging, especially in low- and middle-income countries. Moreover, the observed age and gender differences highlight the importance of incorporating these factors into targeted prevention approaches, which could effectively reduce the disease burden in specific populations and improve overall public health outcomes.

## Data Availability

The data that support the findings of this study are available from: https://vizhub.healthdata.org/gbd-results/. Analysis codes can be obtained by contacting the corresponding author.

## Abbreviations

Nonstandard Abbreviations and Acronyms

ASR: age-standardized rate
BMI: body-mass index
CI: confidence interval
DALY: disability-adjusted life year
EAPC: estimated annual percentage change
GATHER: guidelines for accurate and transparent health estimates reporting
GBD: global burden of disease
IS: ischemic stroke
LOESS: Locally estimated scatterplot smoothing
SDI: socio-demographic index
TMREL: theoretical minimum risk exposure level
UI: uncertainty intervals
WHO: world health organization

## 5 Acknowledgments

This study used publicly available deidentified data accessed from GBD 2021, which are provided by the Institute for Health Metrics and Evaluation.

## 8 Supplemental Material

GATHER Checklist

Tables S1–S3

Figures S1–S2

